# Improving support for family caregivers: a mixed-methods effect evaluation of an organizational intervention

**DOI:** 10.1101/2025.09.08.25335005

**Authors:** Hinke E. Hoffstädt, Arianne Stoppelenburg, Marcella C. Tam, Iris D. Hartog, Leti van Bodegom-Vos, Bart J.A. Mertens, Jenny T. van der Steen, Yvette M. van der Linden

**Affiliations:** Center of Expertise in Palliative Care, Leiden University Medical Center, Leiden, the Netherlands; Biomedical Data Sciences, section Medical Decision Making, Leiden University Medical Center, Leiden, The Netherlands; Medical Statistics, Department of Biomedical Data Sciences, Leiden University Medical Center, Leiden, the Netherlands; Public Health and Primary Care, Leiden University Medical Center, Leiden, The Netherlands; Radboudumc Alzheimer Center and Department of Primary and Community Care, Radboud university medical center, Nijmegen, the Netherlands; Cicely Saunders Institute, King’s College London, UK

**Keywords:** Palliative care, family caregivers, family support, intervention, mixed-methods

## Abstract

**Objectives:** To evaluate the impact of a tailored organizational intervention on the support for family caregivers.

**Methods:** A convergent mixed-methods study was conducted in 17 organizations (6 hospices, 5 home care organizations, 3 nursing homes, 2 hospitals, 1 transmural organization) between November 2021 and August 2023. The intervention comprised a quality improvement trajectory during which each organization conducted a structured workshop to define organization-specific goals to improve their support for family caregivers and to develop an action plan to achieve those goals. The action plan was implemented over one year with intermittent evaluations. Pre and post-intervention surveys were distributed among healthcare professionals (paired) and bereaved family caregivers (non-paired) to assess provided and received support. Data were analyzed with mixed models and regression analyses. Post-intervention focus groups with project team members and final evaluation reports were analyzed with qualitative content analysis.

**Results:** Survey respondents were 97 healthcare professionals (83% nursing staff), 123 family caregivers pre-intervention and 99 family caregivers post-intervention. Only healthcare professionals of home care organizations reported a significant increase in attending to family caregivers’ wellbeing and needs (scale 0-20; β=3.65; 95%CI: 1.33-5.97). Family caregivers’ reports of healthcare professionals attending to their wellbeing and needs did not change (scale 0-2; β=0.17; 95%CI: -0.04-0.38). Across settings, healthcare professionals evaluated the care they provided more positively post-intervention (scale 0-8; β=0.65, 95%CI: 0.38-0.97). In home care, family caregivers also evaluated care more positively (scale 0-8; β=2.12; 95%CI: 0.89-3.34). Four focus groups and 17 evaluation reports indicated improvements at three levels: the support for family caregivers (increased awareness of healthcare professionals, changes in work processes, more structured support), the healthcare team (more skills, confidence, available tools) and the organization (fostering sustainability).

**Significance of results:** A tailored organizational intervention can strengthen the supporting of family caregivers in healthcare organizations.

## Introduction

Support for family caregivers is an integral aspect of palliative care as a life-threatening disease also impacts people close to the patient (World Health Organization 2020). Family caregivers commonly experience physical symptoms such as sleep disturbance and fatigue, and depression, anxiety and distress (Alam et al. 2020). In a longitudinal survey study among advanced cancer patients and family caregivers in the Netherlands, family caregivers’ emotional functioning was lower than that of patients (van Roij et al. 2022). As such, family caregivers should be considered as care recipients alongside the patient (Hudson et al. 2020). Family caregivers also play an important role in the care for the patient. With increasing shortages in healthcare and the ageing population, their role in palliative care provision is becoming even more important (Etkind et al. 2017; Michaeli et al. 2024; PZNL, AHzN, KWF and VPTZ 2020). Therefore, family caregivers being well-supported by healthcare professionals is a prerequisite to ensure high-quality care for the patient as well.

A recent scoping review found that existing clinical guidelines for the support for family caregivers state that healthcare professionals should assess and meet their emotional, practical, physical, social and existential support needs, provide information, involve them in the patient’s care and decision-making, help them in preparing for the patient’s death, and offer bereavement support (Coelho et al. 2025). However, several studies have demonstrated that such support is not structurally embedded in healthcare organizations, for example in the Netherlands and America (Becqué et al. 2021; Hoffstädt et al. 2023; Sabo et al. 2022). Therefore, a change of practice in healthcare organizations is warranted, but this is associated with numerous challenges such as lacking resources, high workload and resistance to change (Geerligs et al. 2018; Williams et al. 2015). Strategies for successful implementation of organizational interventions include flexibility in implementation and adaptation of the intervention to the organization’s existing structures (Collingridge Moore et al. 2020; Powell et al. 2019). To this end, we developed an organizational intervention that could be tailored to the organization’s context to improve support for family caregivers. During the intervention, healthcare professionals engaged in a quality improvement trajectory in which they worked towards achieving organization-specific goals to enhance their current practice of supporting family caregivers. Healthcare professionals took the lead in defining these goals to foster ownership and reduce resistance to change. The aim of this study was to evaluate the impact of this tailored organizational intervention on the support for family caregivers.

## Methods

### Study design

A convergent mixed-methods study design was adopted, with qualitative and quantitative data collected and analyzed independently, followed by their integration to facilitate a holistic understanding of the intervention’s effect (Creswell and Clark 2017). The study was conducted in various healthcare settings between December 2021 and August 2023. Before and after the intervention, surveys were distributed among healthcare professionals (same group pre and post) and recently bereaved family caregivers (different groups pre and post). Additionally, after the intervention, focus groups with participants were conducted and final evaluation reports were written.

This study was part of a larger Dutch study called ‘Support for family caregivers’ (2017-2024; Hoffstädt et al. 2023; Hoffstädt et al. 2024). This substudy was preregistered on the Open Science Framework (OSF) in November 2021 (Stoppelenburg et al. 2021). Some deviations from the preregistration occurred: qualitative data and a survey study among family caregivers were added and some adjustments were made in the outcome measures based on how the healthcare organizations shaped their improvement trajectory. These deviations from the preregistration are described in the ‘measures’ section. The Medical Ethics Committee Leiden Den Haag Delft approved the study (N21.072).

### The intervention and its procedure

In total, 17 organizations (6 hospices, 5 home care organizations, 3 nursing homes, 2 hospitals, 1 transmural organization) implemented the intervention, which comprised a quality improvement trajectory consisting of five steps. First, in each participating organization a project team of 4 to 7 members was assembled, consisting of healthcare professionals from various professions and one member of the management team. One healthcare professional was appointed as the project ambassador to lead the team. Second, the project team and some additional healthcare professionals (the group ranging from 6 to 11 people) conducted the ‘Family caregiver journey’ workshop which was developed as part of the study (Boere 2021). The structured workshop facilitated a discussion on the organization’s current support for family caregivers, its strengths and weaknesses, and a brainstorm on strategies to address shortcomings. Third, based on the workshop’s outcomes, the project teams formulated organization-specific goals and developed an action plan to improve the support for family caregivers. Fourth, the project teams kicked off with an educational session for all healthcare professionals of the involved departments or teams to notify them of the action plan and what is needed from them to achieve the goals. Last, project teams worked towards achieving their goals over the following year. Quarterly evaluation meetings were held to monitor progress. A toolkit was available to facilitate achieving the goals, including brochures with information for healthcare professionals and family caregivers. In each organization, one researcher (MT or HH) maintained contact with the project ambassador to give instructions and advise on the trajectory and to receive updates regarding progress.

### Measures

The surveys were self-developed as no existing instrument covered our purpose. Both surveys addressed demographic characteristics, healthcare professionals’ attention to family caregivers’ wellbeing and needs, more specific support types (e.g. information provision, inquiring after additional support needs) and evaluation of care that was provided or received. Some questions were sourced from existing Dutch family caregiver surveys and adapted to be applicable to healthcare professionals (Hoffstädt et al. 2025; van der Steen et al. 2014). A detailed description of all quantitative outcome measures is provided in Table 1.

**Table 1.**
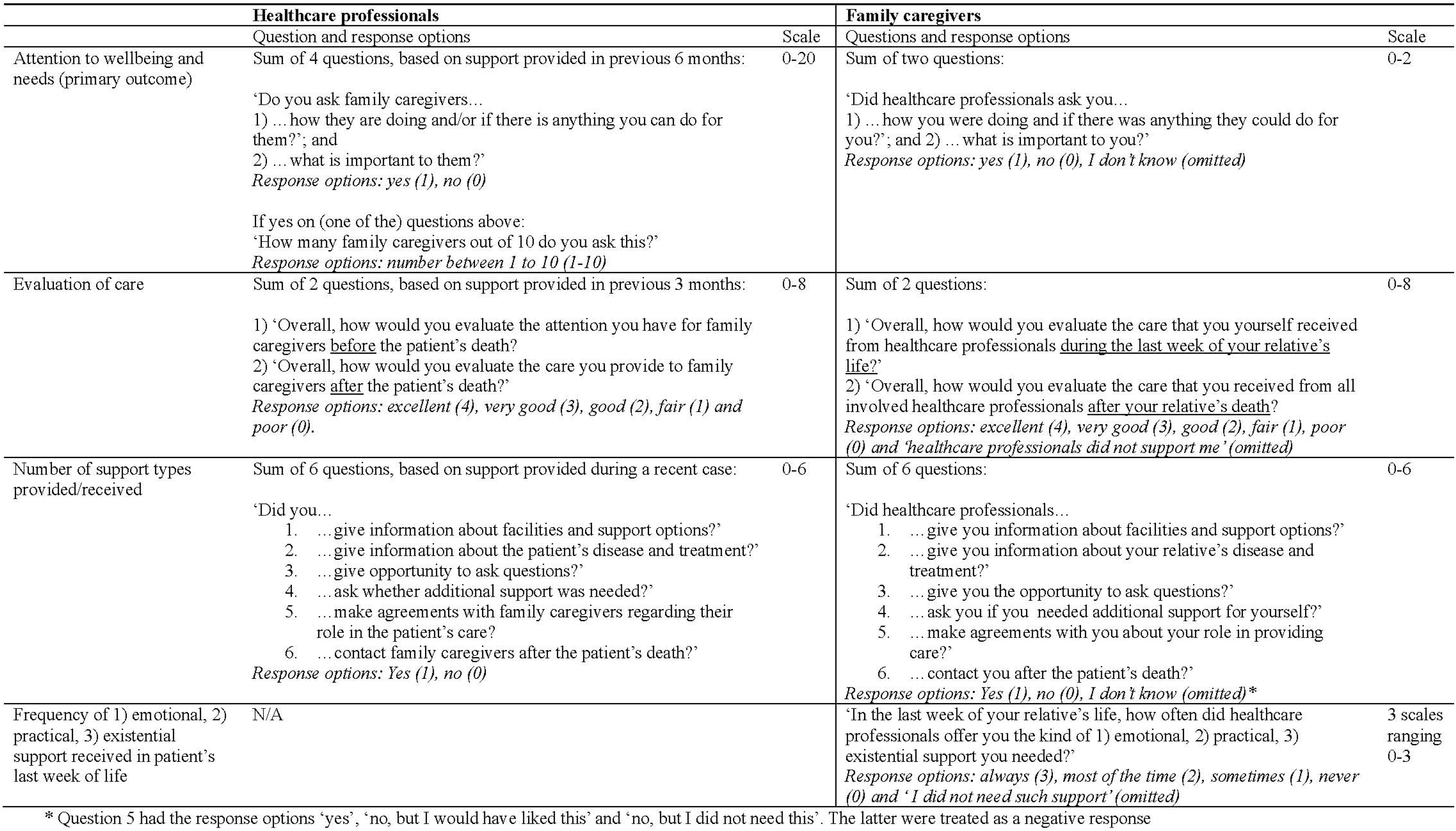
Outcome measures regarding the effect of the ON2 intervention.

#### Primary outcome measure

The primary outcome measure was ‘attention to wellbeing and needs’, which encompassed whether healthcare professionals asked family caregivers how they were doing, if they could do anything for them and what was important to them. For healthcare professionals, a 0-20 scale was computed based on 4 questions. For family caregivers, a 0-2 scale was computed based on 2 questions (Table 1). This slightly deviates from the primary outcome measure preregistered with the OSF, which reads “the number of healthcare professionals who ask at least 80% of family caregivers how they are doing and/or whether they can do something for them.” The adjustment was made to gain a broader understanding of this aspect of care.

#### Secondary outcome measures

The five secondary outcome measures were: 1) evaluation of care; 2) number of support types provided / received; and family caregivers’ reports on the frequency of 3) emotional, 4) practical, and 5) existential support received during the patient’s last week of life (Table 1). The latter four secondary outcome measures were not preregistered with OSF. Furthermore, one secondary outcome measure preregistered with OSF was dropped from analysis: “the needs of healthcare professionals for support in providing pre and post-death care to family caregivers assessed with selected items from the End-of-Life Professional Caregiver Survey.” This outcome measure was dropped as the educational needs included in the survey were rarely targeted by the action plans.

*Evaluation of care:* For both healthcare professionals and family caregivers, a 0-8 scale was computed by summing two questions assessing evaluation of care provided or received *before* and *after* the patient’s death.

*Number of support types provided / received:* For both healthcare professionals and family caregivers, a 0-6 scale was computed by summing 6 items assessing whether a specific support type was provided or received, such as information provision or whether there had been contact after the patient’s death. *Frequency of 1) emotional, 2) practical and 3) existential support received in the patient’s last week of life:* These outcome measures were assessed only among family caregivers. Each outcome comprised one question assessed with a 0-3 scale.

### Data collection

#### Quantitative data

Before the intervention, the research team digitally distributed the survey to the healthcare professionals involved in the project who had regular contact with family caregivers. The family caregiver survey was distributed by postal mail to all family caregivers of patients who had died in the previous 6 months while in care of a participating healthcare team or department. After the intervention, surveys were distributed in the same manner among the engaged healthcare professionals and a new group of recently bereaved family caregivers. Respondents agreed with the use of their data as they completed and returned the survey. Data entry and management were facilitated by Castor EDC (2019).

#### Qualitative data

Project ambassadors completed an evaluation form prior to the final evaluation meeting, which regarded the progress of the set goals, specific changes in work processes, encountered barriers and facilitators, and sustainability of the changes that were made. Furthermore, the involved researcher wrote a report on what was discussed during the final evaluation meeting. Last, online focus groups were conducted after the intervention with project team members of each healthcare setting. The focus groups were guided by a topic list similar to the topics addressed in the final evaluation form. All focus groups were recorded and transcribed clean verbatim.

### Data analyses

#### Quantitative data analysis

First, descriptive statistics were used to present demographic characteristics and the support that was provided or received. Second, for each outcome measure of healthcare professionals, mixed model analyses were performed to assess differences between pre and post-intervention. Participant ID was included as a random factor to account for variability between individuals. Type of healthcare setting and an interaction term between pre-post intervention and healthcare setting were added as fixed factors. If the interaction term was significant, the model was run separately for each healthcare setting. When non-significant, the model was rerun without the interaction effect. As the family caregiver data was unpaired, rather than the participant ID the different healthcare organizations were added as a random factor. Because this accounted only for minor variation, we opted for linear regression analyses. Interaction effects were examined as in the healthcare professionals’ analyses. For scale computation of all outcome measures only complete cases were used.

Both mixed model and regression analyses were initially conducted with robust standard errors using bootstrapping due to heteroscedasticity in the data. However, when analyses were performed separately for each healthcare setting, results were presented with non-robust standard errors as bootstrapping was impossible due to the smaller sample sizes.

All analyses were performed in IBM SPSS (version 29). The 95% confidence intervals were inspected to determine significance.

#### Qualitative data analysis

Focus group transcripts and final evaluation reports were analyzed with conventional qualitative content analysis (Hsieh and Shannon 2005). HH coded the data inductively while being guided by the research aim of exploring the intervention’s impact on the support for family caregivers. Categories were created and discussed with the research group. Analyses were performed in ATLAS.ti (version 24).

#### Integration of quantitative and qualitative data

After both the quantitative and qualitative data were analyzed, the findings were compared to identify overlapping findings and findings unique to one data type. Conclusions informed a narrative in which the data types complemented each other.

## Results

In total, 97 professionals completed the pre and post-survey,* most of whom were nursing staff (83%) and most worked in home care (32%), followed by hospices (25%) and nursing homes (21%; Table 2). Of the family caregivers (Table 3), 129 out of 350 responded to the pre-survey (37% response rate) and 100 out of 216 to the post-survey (46% response rate). Seven of those (pre: n=6; post: n=1) were excluded as open-ended responses reported on experiences with non-participating healthcare organizations. Both before and after the intervention, most family caregivers were the patient’s partner (pre: 39%; post: 46%) or child (pre: 42%; post: 36%). Most were recruited through participating hospices (pre: 42%; post: 64%).

**Table 2.** Characteristics of healthcare professionals at baseline (paired pre-post data of 97 healthcare professionals)

|  | % | n |
| --- | --- | --- |
| Age (mean [SD]) | 48 [13] | 97 |
| Years working at current position (mean [SD]) | 13 [11] | 97 |
| Healthcare setting |  |  |
| Home care | 32 | 31 |
| Hospice | 25 | 24 |
| Nursing home | 21 | 20 |
| Hospital | 16 | 15 |
| Transmural<sup>1</sup> | 7 | 7 |
| Profession |  |  |
| Nurse | 57 | 55 |
| Nurse aide / assistant | 26 | 25 |
| Nurse practitioner | 3 | 3 |
| Spiritual counsellor | 2 | 2 |
| Medical specialist | 1 | 1 |
| Other<sup>2</sup> | 11 | 11 |
| Consultant palliative care |  |  |
| Yes | 5 | 5 |
| No<sup>3</sup> | 95 | 92 |

<sup>1</sup> A healthcare organization that provides nursing care in various healthcare settings.
<sup>2</sup> Living room supervisor / welfare assistant / activities coordinator (n=6), hospice coordinator (n=3), hospice manager (n=1), case manager (n=1).
<sup>3</sup> Three healthcare professionals answered 'no' at baseline and 'yes' at the post-measurement.

**Table 3.** Characteristics of the bereaved family caregivers and their relatives who died (non-paired pre-post data)

| **Bereaved family caregiver** | Pre (N = 123) |  | Post (N = 99) |  |
| --- | --- | --- | --- | --- |
|  | % | n | % | n |
| Sex |  |  |  |  |
| Female | 63 | 77 | 64 | 63 |
| Male | 37 | 46 | 36 | 36 |
| Age (mean [SD]) | 64 [12] | 123 | 65 [13] | 99 |
| Relationship to patient |  |  |  |  |
| Partner | 39 | 48 | 46 | 46 |
| Child | 42 | 52 | 36 | 36 |
| Sibling | 5 | 6 | 8 | 8 |
| Other familial relationship | 10 | 12 | 7 | 7 |
| Other | 4 | 5 | 2 | 2 |
| Country of birth |  |  |  |  |
| The Netherlands | 89 | 110 | 97 | 95 |
| Other | 11 | 13 | 3 | 3 |
| Educational level |  |  |  |  |
| High | 43 | 53 | 48 | 46 |
| Middle | 22 | 27 | 19 | 18 |
| Low | 34 | 42 | 33 | 31 |
| Patient's primary contact person |  |  |  |  |
| Yes | 91 | 104 | 88 | 86 |
| No | 9 | 10 | 12 | 12 |
| Recruited from which type of healthcare organization |  |  |  |  |
| Hospice | 42 | 52 | 64 | 63 |
| Home care organization | 23 | 28 | 12 | 12 |
| Hospital | 16 | 20 | 17 | 17 |
| Nursing home | 13 | 16 | 5 | 5 |
| Transmural organization | 6 | 7 | 2 | 2 |
| **Relative who died** |  |  |  |  |
| Age at death (mean [SD]) | 78 [14] | 122 | 77 [13] | 98 |
| Country of birth |  |  |  |  |
| The Netherlands | 86 | 105 | 92 | 90 |
| Other | 14 | 17 | 8 | 8 |
| Place of death<sup>1</sup> |  |  |  |  |
| Hospice | 45 | 55 | 64 | 63 |
| Home | 24 | 29 | 13 | 13 |
| Hospital | 18 | 22 | 17 | 17 |
| Nursing home | 13 | 16 | 6 | 6 |
| Other | 1 | 1 | 0 | 0 |
| Cause(s) of death (as reported by family caregivers) |  |  |  |  |
| Cancer | 62 | 76 | 68 | 67 |
| Heart failure | 13 | 16 | 9 | 9 |
| Infectious disease | 8 | 10 | 13 | 13 |
| Chronic lung disease | 8 | 10 | 4 | 4 |
| Stroke | 5 | 6 | 2 | 2 |
| Advanced dementia | 5 | 6 | 5 | 5 |
| Age-related decline | 5 | 6 | 3 | 3 |
| Kidney failure | 3 | 4 | 1 | 1 |
| Other<sup>2</sup> | 6 | 7 | 6 | 6 |
| Unknown | 4 | 5 | 7 | 7 |
| Duration of serious health problems before death |  |  |  |  |
| More than two years | 29 | 35 | 28 | 28 |
| More than 6 months, but less than 2 years | 25 | 30 | 19 | 19 |
| Between 3 and 6 months | 18 | 22 | 19 | 19 |
| More than 1 month, but less than 3 months | 13 | 16 | 14 | 14 |
| More than 1 week, but less than 1 month | 12 | 15 | 15 | 15 |
| Less than 1 week | 3 | 4 | 4 | 4 |

<sup>1</sup> In some cases, the patient died in a different setting from where the respondent was recruited (e.g., recruited from home care organization, but the patient died in a hospital)
<sup>2</sup> E.g. ALS, internal bleeding, liver failure, hip fracture.

In total, 22 project team members of 14 of the 17 organizations participated in the focus groups. Most were project ambassadors (n=10), followed by managers (n=8) and other project team members (n=4). An individual interview was conducted with a nursing home manager involved in two of the nursing homes of a single organization who could not attend the focus group.

The qualitative data analysis led to findings regarding the intervention’s impact at three levels that informed the narrative below: the direct impact on the support for family caregivers as provided by healthcare professionals and received by family caregivers, the impact on the healthcare team, and the impact on the healthcare organization. Quantitative findings pertained to the first level. An overview of the descriptive statistics for all survey items is provided in Supplement 1.

### The intervention’s direct impact on the support for family caregivers

Both before and after the intervention, (almost) all healthcare professionals reported to have asked family caregivers how they were doing and/or if they could do anything for them (pre: 98%; post: 100%) and the majority asked at least 80% of family caregivers (pre: 71%; post: 72%). An increase was observed in the number of healthcare professionals reporting to have asked family caregivers what was important to them (pre: 79%; post: 93%; Table 4). The mixed model analysis on ‘attention to wellbeing and needs’ demonstrated a significant increase among healthcare professionals working in home care (β=3.65; 95% CI: 1.33-5.97; Table 5). With regard to family caregivers, more reported to have been asked how they were doing and if anything could be done for them in the post-intervention group (87%) than in the pre-intervention group (74%; Table 4). A smaller increase was observed in family caregivers reporting to have been asked what is important to them (pre: 62%; post: 65%). The linear regression analysis on ‘attention to wellbeing and needs’ received by family caregivers did not demonstrate a significant increase (Table 5). No large increase was observed in reports of having received sufficient support and guidance (pre: 64%; post: 68%). However, after the intervention family caregivers reported more often that sufficient attention was paid to them (pre: 61%; post: 74%).

**Table 4.**
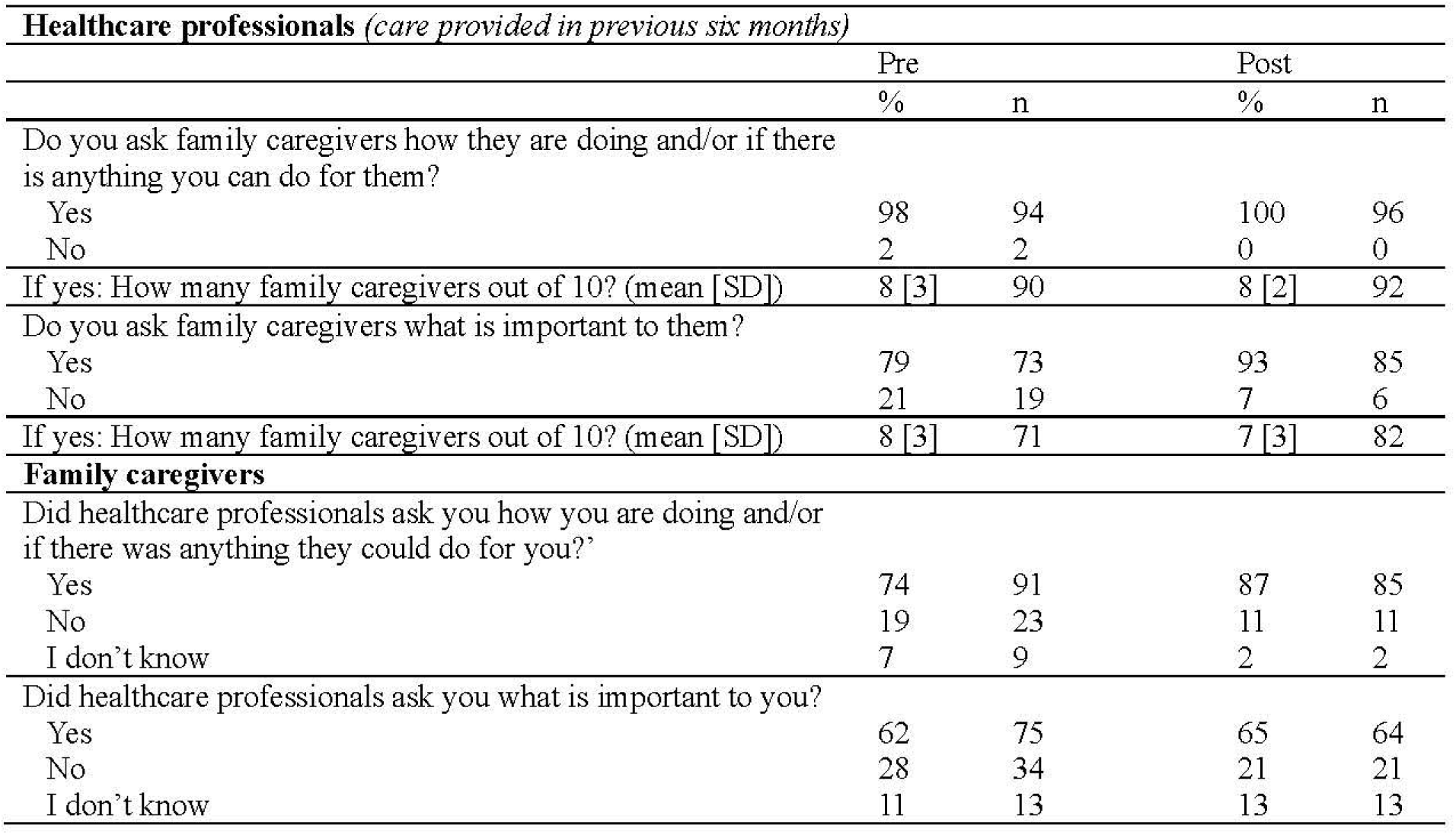
Healthcare professionals’ attention to family caregivers’ wellbeing and needs before and after the intervention.

| **Healthcare professionals** (*care provided in previous six months*) |  |  |  |  |
| --- | --- | --- | --- | --- |
|  | Pre |  | Post |  |
|  | % | n | % | n |
| Do you ask family caregivers how they are doing and/or if there is anything you can do for them? |  |  |  |  |
| Yes | 98 | 94 | 100 | 96 |
| No | 2 | 2 | 0 | 0 |
| If yes: How many family caregivers out of 10? (mean [SD]) | 8 [3] | 90 | 8 [2] | 92 |
| Do you ask family caregivers what is important to them? |  |  |  |  |
| Yes | 79 | 73 | 93 | 85 |
| No | 21 | 19 | 7 | 6 |
| If yes: How many family caregivers out of 10? (mean [SD]) | 8 [3] | 71 | 7 [3] | 82 |
| **Family caregivers** |  |  |  |  |
| Did healthcare professionals ask you how you are doing and/or if there was anything they could do for you? |  |  |  |  |
| Yes | 74 | 91 | 87 | 85 |
| No | 19 | 23 | 11 | 11 |
| I don't know | 7 | 9 | 2 | 2 |
| Did healthcare professionals ask you what is important to you? |  |  |  |  |
| Yes | 62 | 75 | 65 | 64 |
| No | 28 | 34 | 21 | 21 |
| I don't know | 11 | 13 | 13 | 13 |

**Table 5.** Intervention effects on care provided and care received - outcomes of mixed model and linear regression analyses.

| Healthcare professionals (mixed model analyses) |  |  | Family caregivers (linear regression models) |  |  |
| --- | --- | --- | --- | --- | --- |
| | $\beta^1$ | [95% CI]<sup>1</sup> | | $\beta$ | [95% CI] |
| Attention to needs and wellbeing*<sup>2</sup> |  |  | Attention to needs and wellbeing | 0.17 | [-0.04 – 0.38] |
| Hospital | 0.93 | [-1.32 – 3.19] |  |  |  |
| Nursing home | -1.40 | [-3.93 – 1.13] |  |  |  |
| Hospice | 2.09 | [-0.17 – 4.35] |  |  |  |
| **Home care** | **3.65** | **[1.33 – 5.97]** |  |  |  |
| Other | 1.38 | [-0.82 – 3.58] |  |  |  |
| **Number of support types provided** | **0.67** | **[0.12 – 1.27]** | Number of support types received | -0.16 | [-0.66 – 0.34] |
| **Evaluation of care** | **0.65** | **[0.38 – 0.97]** | Evaluation of care* |  |  |
|  |  |  | Hospital | 0.40 | [-1.29 – 2.09] |
|  |  |  | Nursing home | 0.20 | [-3.50 – 3.90] |
|  |  |  | Hospice | 0.16 | [-0.54 – 0.86] |
|  |  |  | **Home care** | **2.12** | **[0.89 – 3.34]** |
|  |  |  | Other | 3.00 | [-4.12 – 10.12] |
|  |  |  | Frequency emotional support* |  |  |
|  |  |  | Hospital | -0.55 | [-1.24 – 0.13] |
|  |  |  | Nursing home | 0.05 | [-1.43 – 1.52] |
|  |  |  | Hospice | 0.22 | [-0.14 – 0.58] |
|  |  |  | Home care | 0.85 | [-0.07 – 1.76] |
|  |  |  | Other | 1.20 | [-0.91 – 3.31] |
|  |  |  | Frequency practical support | -0.09 | [-0.48 – 0.28] |
|  |  |  | Frequency existential support | 0.05 | [-0.40 – 0.52] |

<sup>1</sup> $\beta$ = beta coefficient, 95% CI = 95% confidence interval
<sup>2</sup> In the models with an asterisk a significant interaction term implied a difference between pre- and post-intervention assessments and differences by type of healthcare setting. Therefore, these models were run separately for healthcare settings.

Qualitative data also implied improvement on healthcare professionals’ attention to family caregivers wellbeing and needs as, across all organizations, an increased awareness of healthcare professionals of the importance of doing so was reported. Project team members mentioned that healthcare professionals more frequently engaged in informal conversations with family caregivers. In some cases, the intervention also brought about *“that there is now more focus on family caregivers from the intake, instead of that being something that has to grow over time.”* (researcher’s evaluation report; home care organization).

#### New and improved actions of healthcare professionals to support family caregivers

The quantitative data showed increases in healthcare professionals’ reports on whether they provided specific types of support during a recent case (Figure 1). The largest increase was found in making agreements with family caregivers about their role as co-caregiver (pre: 60%; post: 79%) and information provision about specific facilities and support options (pre: 51%; post: 65%). The mixed model analysis on ‘number of support types provided’ demonstrated a significant increase (β=0.67, 95% CI: 0.12-1.27; Table 5). Among the family caregivers, no appreciable increases were observed in specific support types that were received before the patient’s death. Increase were observed in having had contact with healthcare professionals after the patient’s death (pre: 63%; post: 73%) and a follow-up conversation (pre: 33%; post: 46%; Figure 1). The linear regression analysis on ‘number of support types received’ did not demonstrate a significant increase. Nor did the linear regression analyses on the frequency in which family caregivers reported having received the kind of emotional, existential and practical support they wanted (Table 5, Table 6).

**Figure 1.**
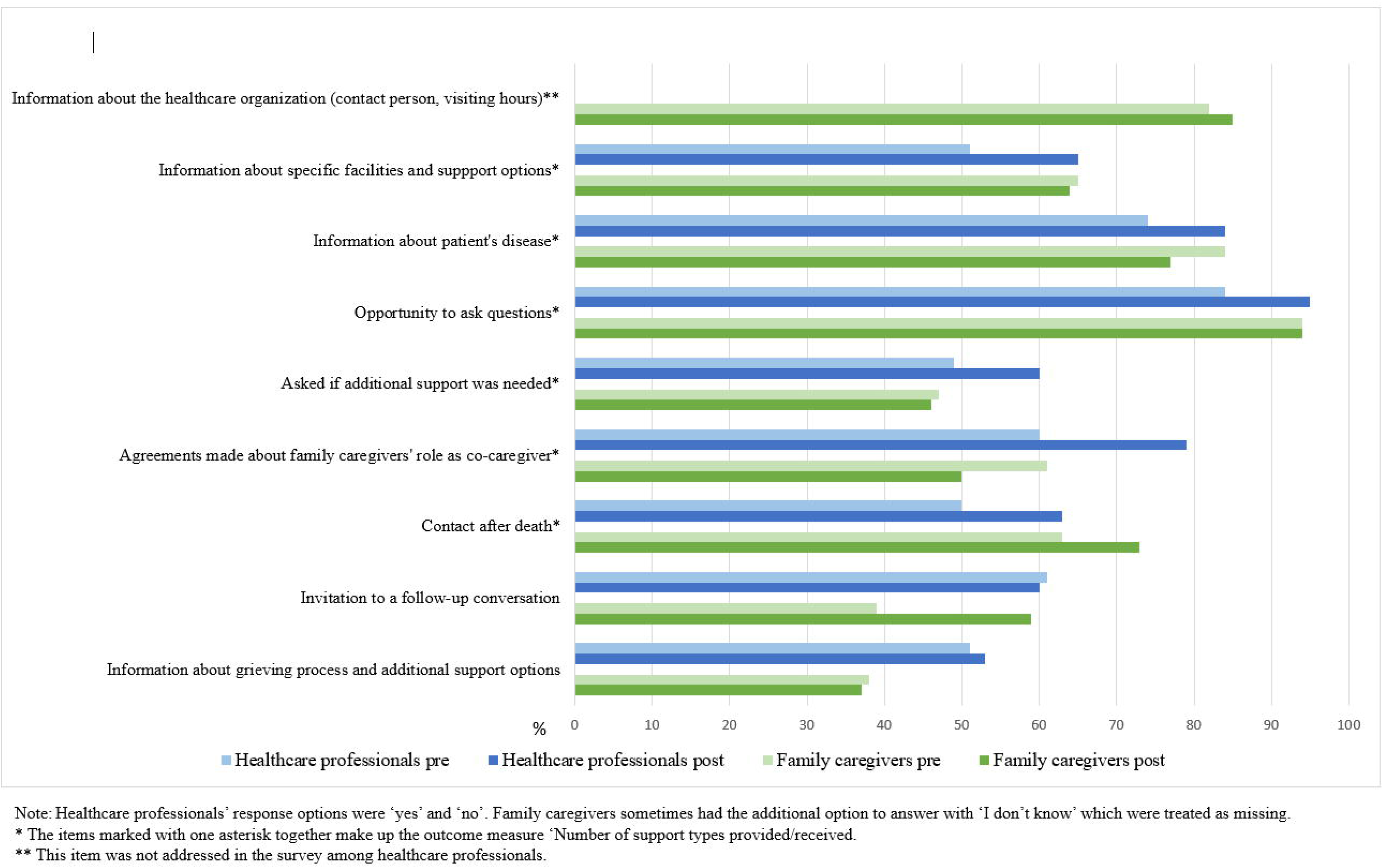
Specfic care provided by healthcare professionals and recieved by family caregivers

**Table 6.** Frequency of emotional, practical and existential support that was received by family caregivers before and after the intervention.

| Were you in need of... | ... **emotional** support? |  |  |  | ... **practical** support? |  |  |  | ... **existential** support? |  |  |  |
| --- | --- | --- | --- | --- | --- | --- | --- | --- | --- | --- | --- | --- |
|  | Pre |  | Post |  | Pre |  | Post |  | Pre |  | Post |  |
|  | % | n | % | n | % | n | % | n | % | n | % | n |
| Yes | 76 | 93 | 80 | 77 | 73 | 88 | 60 | 56 | 55 | 65 | 41 | 39 |
| No | 24 | 29 | 20 | 19 | 27 | 32 | 40 | 37 | 45 | 54 | 59 | 57 |
| If in need: How often, in the last week of life of your relative, did the healthcare professionals offer the kind of... |  |  |  |  |  |  |  |  |  |  |  |  |
|  | ... **emotional** support you wanted? |  |  |  | ... **practical** support you wanted? |  |  |  | ... **existential** support you wanted? |  |  |  |
|  | Pre (n=93) |  | Post (n=77) |  | Pre (n=88) |  | Post (n=56) |  | Pre (n=65) |  | Post (n=39) |  |
|  | % | n | % | n | % | n | % | n | % | n | % | n |
| Always | 31 | 29 | 39 | 30 | 38 | 33 | 29 | 16 | 25 | 16 | 23 | 9 |
| Most of the time | 31 | 29 | 30 | 23 | 20 | 18 | 27 | 15 | 17 | 11 | 21 | 8 |
| Sometimes | 25 | 23 | 29 | 22 | 25 | 22 | 25 | 14 | 28 | 18 | 31 | 12 |
| Never | 13 | 12 | 3 | 2 | 17 | 15 | 20 | 11 | 31 | 20 | 26 | 10 |

The qualitative data illustrated several and diverse changes in work processes of healthcare professionals. In some cases, this entailed improving existing practices, while in other cases, new practices were introduced. An overview of new and improved actions is presented in the first box of Figure 2. The adjusted work processes related to providing emotional support and information, family caregivers’ involvement in the patient’s care, additional support and support after the patient’s death. The intervention also contributed to the support for family caregivers becoming more structured: *“On the one hand, we did a lot of things that were good, but not in a very structured way. Everyone did it in their own way. And we have been able to really improve this.”* (project ambassador during focus group; nursing home).

**Figure 2.**
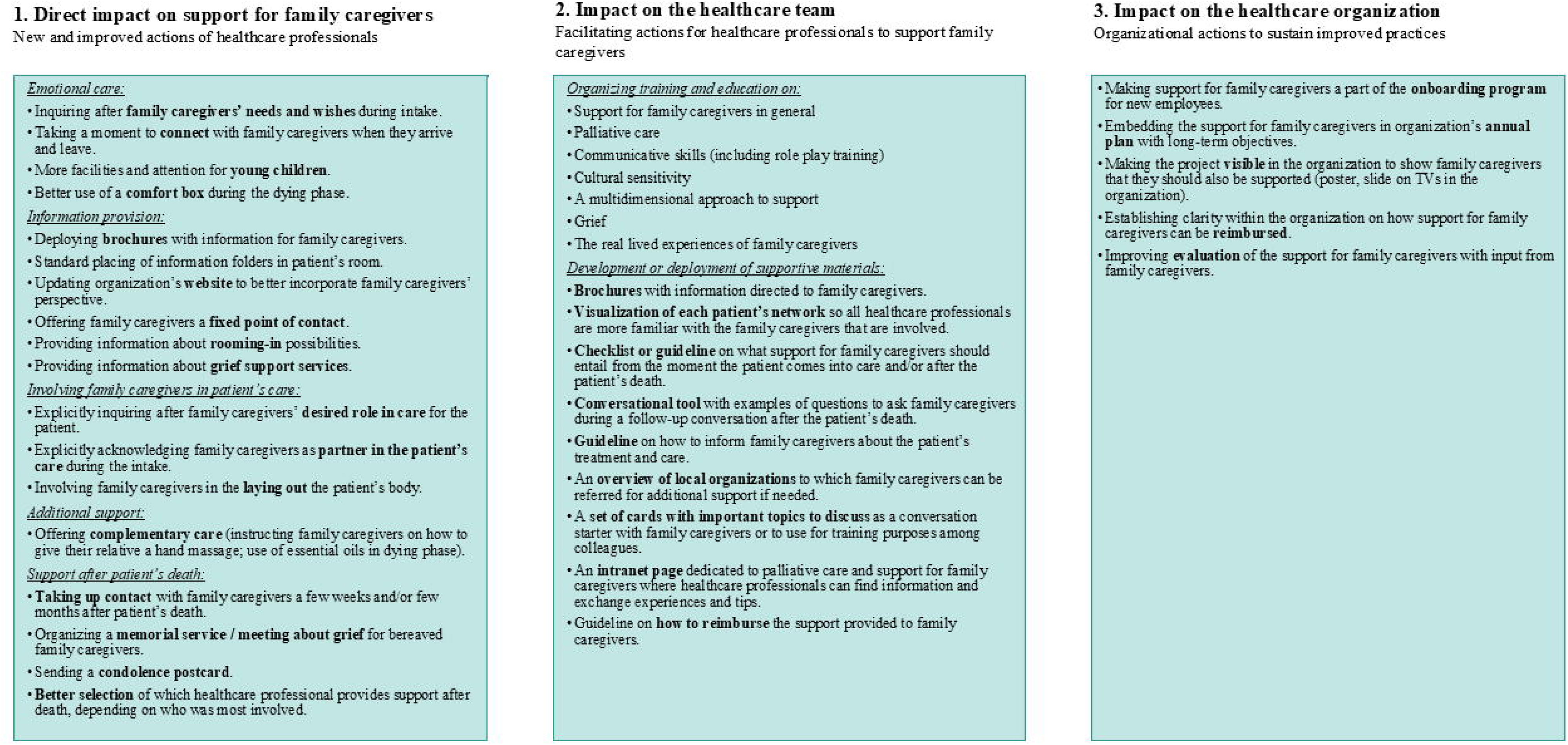
Changesin practices related to support for family caregivers asdemonstrated by qualitative data.

#### Evaluation of care for family caregivers

After the intervention, more healthcare professionals evaluated the attention they had for family caregivers before the patient’s death in the previous three months as ‘excellent’ or ‘very good’ than before (pre: 28%; post: 38%). This was also the case for their evaluation of the care they provided after the patient’s death (pre: 10%; post: 23%; Table 7). The mixed model analysis demonstrated a significant increase in their overall evaluation of the care they provided (β=0.65; 95% CI: 0.38-0.97; Table 5). With regard to the family caregivers, those in the post-intervention group evaluated the care they received in their relative’s last week of life more commonly as ‘excellent’ or ‘very good’ than those in the pre-intervention group (pre: 52%; post: 66%). This was also the case for their evaluation of care after the patient’s death (pre: 39%; post: 53%; Table 7). The linear regression analysis on their overall evaluation demonstrated a significant increase only among those who received care from home care organizations (β=2.12; 95% CI: 0.89-3.34; Table 5).

**Table 7.**
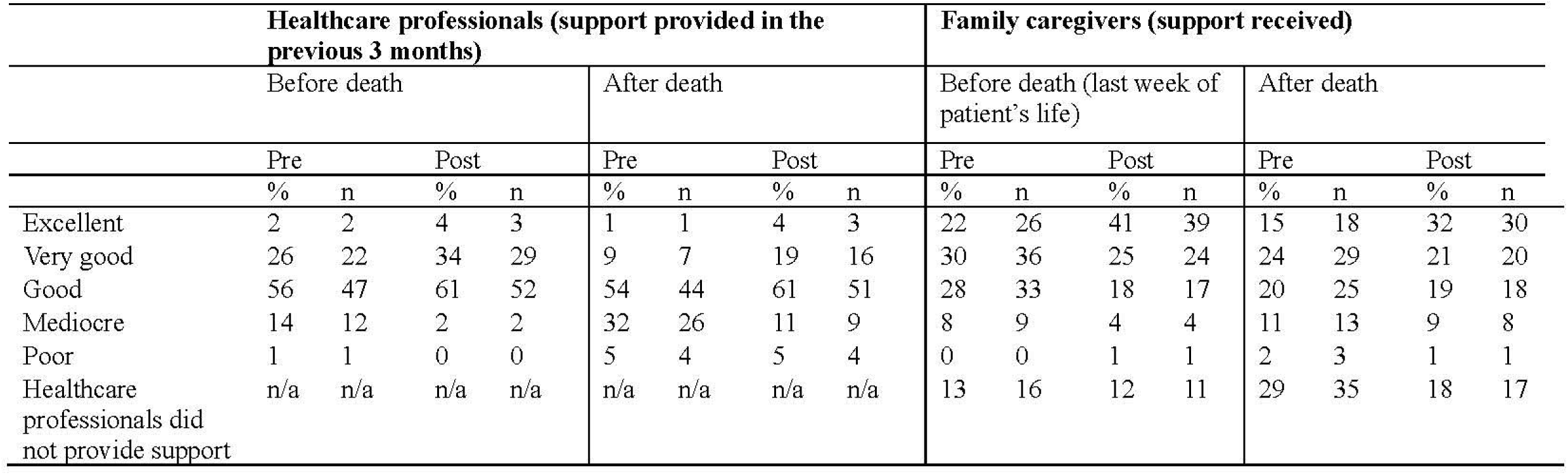
Evaluation of care provided by healthcare professionals and received by family caregivers before and after the intervention.

In an evaluation report was mentioned that *“family caregivers were more satisfied and felt seen”* (project ambassador’s evaluation report, transmural organization). Elsewhere was mentioned that, since the intervention, “*more positive reactions and expressions of gratitude were expressed [by family caregivers] at the participating hospital department compared to other departments”* (researcher’s evaluation report, hospital).

### The intervention’s impact on the healthcare team

#### Support for family caregivers as a part of the workplace culture

Multiple project team members noted that due to the intervention and the increased awareness it brought about, the support for family caregivers became more commonplace and embedded in the workplace culture. In some cases, family caregivers were more often mentioned in electronic health records and they became more often a topic of discussion during team meetings: *“Every healthcare professional is much more aware of the importance of providing good care for the family caregivers. This is increasingly considered as a given.”* (manager’s evaluation report; hospice).

#### Facilitating actions for healthcare professionals

The intervention also helped to better equip healthcare professionals and increase their confidence in supporting family caregivers. In all organizations, one or more educational sessions were organized. Some project teams also worked on creating guidelines or checklists on what support for family caregiver should entail in general or on more specific elements. In some cases, it was explicitly mentioned that such educational sessions and availability of tools contributed to *“confidence among employees in terms of knowledge and skills”* (project ambassador’s evaluation report; home care organization). An overview of those facilitating actions is provided in the second box of Figure 2.

#### Positive spillover effects

In a few instances, the intervention brought about positive spillover effects for healthcare teams. For example, the intervention contributed to better collaboration with other disciplines or among colleagues. Personal experiences came up in the discussions during the journey workshop or during the educational sessions, which contributed to the team becoming more close-knit. This also resulted in more insight in personal experiences of colleagues and strengths and weaknesses with regard to support for family caregivers, which helped to see how team members might complement each other: *“You have your specializations within your team. And you can also kind of look at, what kind of family caregiver is in front of you and who would be compatible. And then to start with ‘How would you prefer to die yourself?’. During the training they had to tell each other about that, so then you know: oh those two [healthcare professional and family caregiver] are a match for such-and-such reason. (…) Then you can say, gosh, you know, that’s really your thing, or you’re good at that, would you please take the time to do that, because you are just better suited for it.”* (project ambassador during focus group; nursing home).

Furthermore, the intervention and the related discussions made healthcare professionals more aware of the potential impact of a patient’s death on their own wellbeing, leading them to pay more attention to each other when a patient died.

### The intervention’s impact on the healthcare organization

In some cases, the intervention also brought about changes at the organizational level to help sustain the new practices and to further improve the support for family caregivers. An overview of all actions at the organizational level is provided in the third box of Figure 2. Examples are incorporating the support for family caregivers in the annual plan of the healthcare organization and making the support for family caregivers a part of the onboarding program of new employees. Also, more clarity on reimbursement of support for (bereaved) family caregivers was established in some organizations.

### Unachieved goals and barriers

Not all goals from the action plans were fully achieved in all healthcare organizations during the year of implementation. In some cases, the actions described in the action plans proved difficult to make routine practice in one year time. Examples are keeping track of positive experiences with the patient while they were in care to give this to family caregivers after the patient’s death, standard use of a newly developed checklist on support for family caregivers, setting up a room for family caregivers to retreat, standard distribution of brochures to family caregivers, involving family caregivers in multidisciplinary team meetings, and structurally taking up contact after the patient’s death. Frequently mentioned reasons for (partly) unachieved goals were staff shortages and high workload which made it difficult to find the time to work on the action plan and to keep the project at the forefront of healthcare professionals’ minds. Other reasons were a lack of confidence of healthcare professionals to support family caregivers, a lack of managerial support, the absence of a strong project leader who had sufficient time for the project, and large ongoing organizational changes such as mergers or expansions.

## Discussion

In this study, a mixed-methods effect evaluation was conducted of a tailored organizational intervention on the support for family caregivers of patients with life-threatening diseases. Quantitative findings were mixed with statistical analyses indicating improved support for family caregivers, especially in healthcare professionals’ reports, but this pattern not being consistent across all types of support and all outcome measures. Qualitative findings showed an increased awareness of healthcare professionals of the importance of supporting family caregivers and numerous improvements in healthcare professionals’ work processes related to emotional care, information provision, family caregivers’ involvement in the patient’s care, and support after the patient’s death.

Family caregivers’ reports of support that was received after the patient’s death showed more improvement compared to the support before the patient’s death. This may be due to recollections of support after death being more distinct as it consists of fewer interactions with fewer healthcare professionals (Boven et al. 2022; Coelho et al. 2025). Furthermore, in some organizations support after death was (re)introduced as part of the intervention, whereas support before death was already present to a certain extent prior to the intervention. Therefore, it can be understood that the contrast between the pre- and post-measurement reports of support received after the patient’s death was more pronounced.

The mixed quantitative findings in this study may in part be attributed to the challenges of quantitatively assessing the effectiveness of a complex intervention that allows a great degree of flexibility in implementation. It has been emphasized in multiple studies that capturing the effect of tailored, context-specific interventions with quantitative measures generally falls short (Datta and Petticrew 2013; Jansen et al. 2006; Skivington et al. 2021). Although the core components of the present intervention were standardized, considerable variation existed in the specific support types that were targeted by each participating healthcare organization. Therefore, standardized quantitative outcome measures across all organizations may not fully capture the impact of the intervention (Skivington et al. 2021).

Taking all data into consideration, it can be concluded that the intervention had a positive impact on the support for family caregivers. However, not all goals were (fully) achieved. This is not surprising as literature has reported an average of 10% of improvement on main targets after implementation of interventions aiming to change clinical practice (Grol and Grimshaw 2003). Various challenges to implementation are known, such as low resources and high workload of healthcare professionals (Geerligs et al. 2018; Parmar et al. 2022). Taking this into consideration, a year of implementation may be insufficient to sustainably improve various elements of support for family caregivers. The intervention may need to be integrated into the organization’s culture, which can be achieved by reevaluating the support for family caregivers after a year of implementation and establishing a new action plan with refined (partly) unmet goals and/or deciding on new areas for improvement. As such, support for family caregivers can be improved in phases, depending on what is feasible within the organization at that moment in time.

The intervention has several advantages. First, the intervention’s flexibility makes it suitable for any healthcare organization that provides care for patients with life-threatening diseases. Depending on what is feasible within their organization, project teams can decide on the extensiveness of their goals and the scale of implementation. However, certain preconditions should always be met, the most important of which are managerial support, allocated time for project team members to manage the trajectory, and no large reorganizations or other ongoing projects (Cowie et al. 2020; Geerligs et al. 2018; Mathieson et al. 2019). Second, as the healthcare professionals themselves are in charge of the action plan’s content, a sense of ownership is facilitated which increases chances of success and sustainability of the intervention’s effects (Cowie et al. 2020). Last, as the intervention targets organizational structures and healthcare teams, it is complementary to existing interventions that directly target family caregivers through, for instance, structured needs assessment, psycho-education, skills training, facilitating their self-care or organizing family meetings (Aoun et al. 2018; Becqué et al. 2019; Becqué et al. 2023; Gonella et al. 2022; Theißen et al. 2024). The current intervention offers a framework within which such interventions can be integrated if they align the action plan.

### Strengths and limitations

A strength of this study is its mixing of quantitative and qualitative methods, enabling a deep understanding of the intervention’s effect on support for family caregivers. However, the study also has limitations. First, uncertainty exists on whether family caregivers’ survey responses were solely based on their experiences with the participating healthcare organization as they may also have experienced care from non-participating healthcare organizations (Wolf et al. 2021). Second, the vast majority of participating healthcare professionals were nursing staff limiting the generalizability of the findings to other professions. Third, as a suitable survey did not exist, the survey that was used was not validated. Last, the small sample sizes within each healthcare setting limited the statistical power of the mixed model and linear regression analyses.

### Future research

Future research is necessary to further explore the intervention’s impact on the support for family caregivers. First, uncertainty exists regarding the sustainability of the improvements that have been implemented. The long-term effect of the intervention on the support for family caregivers should be investigated, including the barriers and facilitators to its sustainment. Second, research on a larger scale is needed to explore whether the intervention’s effectiveness differs for different healthcare settings or professions. Third, future research may target volunteers as they also have an important role in palliative care provision (Bloomer and Walshe 2020).

## Conclusion

In conclusion, a tailored intervention in healthcare organizations improves the support provided to family caregivers. Due to its flexibility the intervention can be adopted across all kinds of healthcare organizations. However, preconditions such as managerial support and sufficient time for project teams should be met. Future research should investigate the long-term effect of the intervention and the barriers and facilitators to sustaining it.

## Data Availability

All data produced in the present study are available upon reasonable request to the authors.

## Acknowledgements

We would like to thank the healthcare organizations who participated in our study, especially the project team members who carried out the intervention, and the family caregivers who participated in the survey study. We are indebted to Willemijn Boere, MSc, and Dr.ir. Marieke Sonneveld of the Delft University of Technology for the development of the ‘Family caregiver journey’ workshop.

## Footnotes

* The survey was initially distributed to volunteers working at hospices as well, which was later deemed unsuitable as the survey primarily focused on healthcare professionals’ responsibilities. Therefore, we are unable to report a response rate exclusively for healthcare professionals.

